# A Novel Carotid Artery Stent System Integrating Balloon, Stent, and Embolic Protection Device Retrieval Catheter

**DOI:** 10.64898/2025.12.04.25341672

**Authors:** Shengli Shen, Shijie Li, Tao Liu, Cheng Zhang, Hongzhou Duan

## Abstract

**Background:** Carotid artery stenosis is a common cause of ischemic stroke, and carotid artery stenting is an important treatment for carotid artery stenosis. However, current carotid stents face issues such as cumbersome operation procedures and frequent difficulties in retrieving embolic protection devices which can lead to complications.

**Objectives:** This study evaluated the safety and efficacy of a novel carotid artery stent system which integrates balloon, stent and embolic protection device retrieval catheter.

**Methods:** A total of ten Bama miniature pigs were used to establish carotid artery stenosis models. Angiography, optical coherence tomography (OCT) and Doppler ultrasound were performed to assess the degree of stenosis improvement and in-stent restenosis. HE staining was utilized to detect intimal hyperplasia.

**Results:** All stent implantation procedures were successful in 10 pigs. The stenosis degree was significantly improved immediately after stent implantation with residual stenosis <5% and follow-up exams at 5 months postoperatively showed no significant in-stent restenosis and intimal hyperplasia.

**Conclusions:** The novel carotid artery stent system appears safe and can effectively improve the stenosis degree of the carotid artery.

## Background

Stroke is currently the leading cause of death and disability in China^1^, with Ischemic stroke accounting for approximately 69.6–70.8% of all strokes, and carotid artery stenosis is an important cause of ischemic stroke^2^. According to the results of China’s national public health service project—*China Stroke Prevention and Control Report 2019*—the detection rate of high-risk stroke populations among individuals aged 40 and above is 19.84%, and 1.47% of this high-risk population has severe carotid artery stenosis^3^. Carotid artery stenting (CAS), with its advantages of minimal invasiveness, short operation time, reduced average hospital stay, and better patient experience, has increasingly become the preferred treatment option for patients^4^. However, the current surgical procedure of carotid artery stenting is cumbersome, which could be improved.

The standard CAS procedure currently is as follows: 1.Perform angiography to determine the lesion. 2.Measure lesion length and proximal/distal vessel diameters to select appropriate embolic protection devices (EPD), balloons, and stents. 3.Advance the guiding catheter into the affected common carotid artery, 2–3 cm proximal to the lesion. 4.Advance the EPD gently through the lesion, deploy it, and confirm proper expansion under fluoroscopy. 5.Advance the pre-selected balloon to the lesion, inflate to “standard pressure”, deflate, and withdraw the balloon. 6.Advance the self-expanding stent slowly to cover the entire lesion and deploy it. 7.Withdraw the stent delivery system and perform angiography. If significant residual stenosis is observed, perform balloon post-dilation, and then withdraw the balloon. 8.Advance the EPD retrieval sheath gently through the stent to retrieve the EPD. (See Video Supplement 1.)

From the above, there are several critical challenges in current CAS procedures. Firstly, multiple devices, including EPD, pre-dilation balloons, self-expanding stents, post-dilation balloons, and EPD retrieval sheaths, must be sequentially delivered into or removed from the blood vessel, prolonging procedure time, increasing intraoperative X-ray exposure, and raising the risk of vascular injury. It is generally agreed upon that the longer and more complex the procedure, the higher the risk of adverse events like periprocedural stroke^5^. Secondly, in some cases with extremely tortuous blood vessels or calcified plaques, stent struts of some open-cell stents may protrude into the vascular lumen, obstructing the retrieval sheath, leading to retrieval difficulties^6^. Currently, the first issue is addressed mainly through operator experience and meticulous technique, which requires a relatively long learning curve. For the second issue, skilled operators may attempt repeated balloon dilation, adjusting guiding catheter rotation, or using custom-shaped catheters (e.g., 5F MPA1) to retrieve the EPD^7^. In extreme cases, surgical incision may be required to remove the EPD^8^. Therefore, innovating and improving existing stent systems is an urgent clinical need.

Through extensive clinical experience in CAS procedures, our research team designed a novel carotid stent system integrating balloon dilation, stent deployment, and EPD retrieval into a single device. A patent has been granted (Patent No. ZL 202220441399.7). All components and the whole system of this novel carotid stent system had undergone corresponding in vitro testing. In this study, we intend to validate its safety and efficacy in a porcine carotid artery stenosis model.

## Methods

### Introduction to the Novel Carotid Stent System

The novel carotid artery stent system consists of three components at its distal end: a catheter-like structure at the tip (for EPD retrieval), a balloon in the middle section, and a proximal stent system where the self-expanding stent is enclosed in a sheath. The catheter, balloon and stent share a common lumen, with a proximal exit port to facilitate rapid exchange of the micro-guidewire. The distal catheter functions to retrieve the EPD. The middle balloon can be used for both pre-dilation of the stenotic segment and post-dilation of residual stenosis after stent deployment. The balloon lumen is connected proximally and can be inflated or deflated by connecting to a pressure pump. The proximal stent is enclosed within the sheath and can be precisely deployed at the stenotic site. See Figure 1.

**Figure 1.**
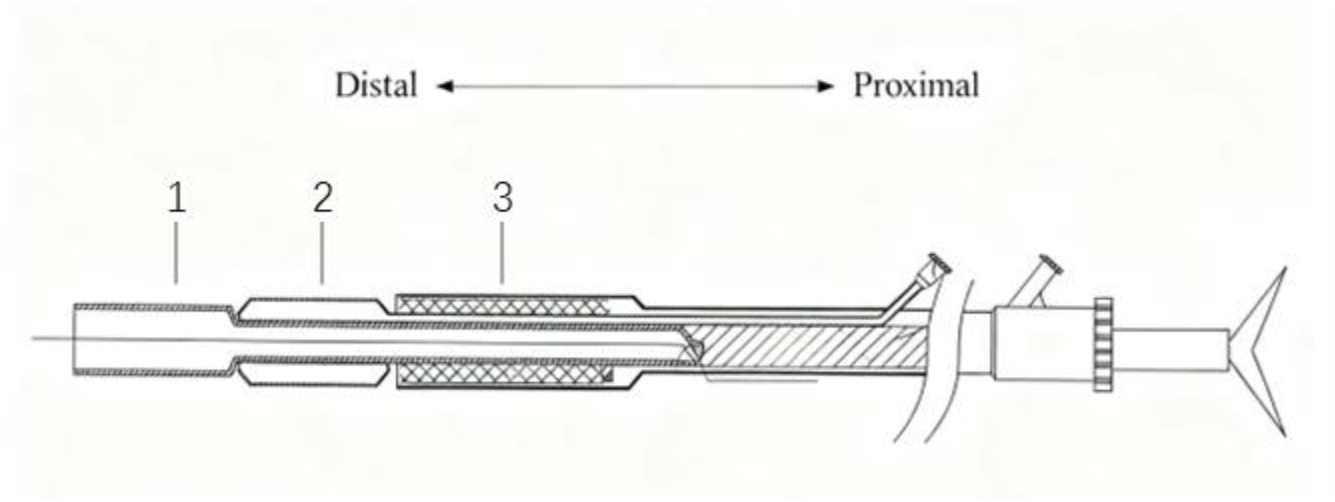
Schematic diagram of the novel carotid stent system. 1 the EPD retrieval catheter, 2 the balloon, 3 the self-expanding stent.

### Operation Procedure

The processes of angiography, measurement of the stenotic segment and EPD deployment are the same as those in conventional CAS. After deploying the EPD distal to the stenosis, the novel carotid artery stent system is rapidly exchanged via the micro-guidewire at the proximal end of the EPD. Advance the novel carotid artery stent system to position the middle balloon at the stenotic site. Inflate the balloon via the proximal pressure pump for pre-dilation, then deflate the balloon and advance the system directly to position and precisely deploy the stent at the stenosis. If significant residual stenosis remains, retract the system to position the balloon to the site of the most severe residual stenosis for post-dilation. Then advance the system to retrieve the EPD using the distal catheter. (See Video Supplement 2.)

### In vitro experiment

All components and the whole system of this novel carotid stent system underwent corresponding in vitro testing. Balloon tests included: leakage test, fatigue test, burst test, and compliance test. Stent tests included: radial force, stent AF point, and dimensional inspection. System tests included: friction test and tensile strength test.

### Experimental Animals

This study was approved by the Experimental Animal Ethics Committee of Peking University First Hospital (No. J2024059). All procedures were performed in accordance with the ARRIVE guidelines (Animal Research: Reporting of In Vivo Experiments). The experimental animals were 10 male Bama miniature pigs, weighing approximately 20-30kg. They were housed in facilities with temperature maintained at 20-25°C, relative humidity 50%-70%, and adequate ventilation. All animals were fed a high-fat, high-cholesterol diet. The high-fat diet was prepared by mixing 6% cholesterol, 12% peanut oil and 82% standard pellet feed.

### Establishment of Carotid Artery Stenosis Model via Fat Compression Method

All surgical and interventional procedures were performed under anesthesia after 12 hours of fasting. After intramuscular ketamine anesthesia, the pigs were immobilized and intravenous access was established. Anesthesia was induced with intravenous propofol (4mg/kg) and midazolam (0.1-0.5mg/kg). After induction, the pigs were intubated and connected to a ventilator with tidal volume set at 170ml. Anesthesia was maintained with a mixture of oxygen and isoflurane (1.0%-2.0%).

After successful anesthesia, the Bama miniature pigs were fixed supine on the operating table. A neck incision was made to expose the right carotid artery. A small amount of subcutaneous fat tissue was harvested, wrapped with appropriately sized sterile glove material, and placed circumferentially around the carotid artery to compress and narrow it. When the degree of carotid artery stenosis reached severe stenosis (≥70%), the glove material was fixed with 4-0 prolene sutures. The subcutaneous tissue and skin were sutured layer by layer. Subsequent angiography confirmed the degree of stenosis. The angiography procedure is described below. After surgery, the high-fat and high-cholesterol diet was continued.

### Implantation of the Novel Carotid Stent

After general anesthesia, the experimental Bama miniature pigs underwent femoral artery puncture and 6F sheath placement. The guiding catheter was positioned in the right carotid artery, and a roadmap was created while recording the carotid stenosis rate. Then the micro-guidewire was advanced through the carotid stenosis into the distal ascending pharyngeal artery, and the EPD was positioned and deployed distal to the stenosis under guidance of the micro-guidewire. The novel carotid stent system was guided by the micro-guidewire to the carotid stenosis site. Balloon dilation was first performed in roadmap mode, and the stenosis improvement rate was recorded via angiography. After balloon dilation, the stent was advanced to the stenotic site and precisely deployed, followed by angiography to record the improvement in stenosis. If the residual stenosis remains severe, retract the balloon to the stenotic site for postdilation. After that, the stent system was further advanced distally to retrieve the EPD. The entire device and guiding catheter were withdrawn, and the puncture site was sutured.

### Tissue Sampling and Evaluation of Stent Efficacy

Optical coherence tomography (OCT) and carotid artery Doppler ultrasound were performed on the miniature pigs to assess the conditions of the local vascular wall and stenosis before and immediately after stent implantation respectively. Three miniature pigs were euthanized immediately after stent implantation by exsanguination, and carotid artery specimens were collected for pathological examination to observe vascular wall and stent conditions. During specimen collection, the stented carotid artery was exposed through a neck incision, and a 4cm segment of the vascular wall at the stenotic site was harvested. The vascular specimens were fixed in 10% formaldehyde solution, routinely embedded in paraffin, sectioned at about 5μm thickness, stained with HE, and observed under light microscopy. The remaining miniature pigs continued on the high-fat, high-cholesterol diet for 5 months after stent implantation, then underwent repeat OCT, carotid ultrasound and angiography using the same methods, followed by euthanasia, specimen collection and HE staining using the same protocols.

### Statistical analysis

The Friedman test was used to analyze the differences in the degree of carotid artery stenosis before stent implantation, immediately after stenting, and at 5-month follow-up postoperatively. If the difference was statistically significant, the Wilcoxon signed-rank test with Bonferroni correction was performed for post-hoc pairwise comparisons. Data was analyzed using IBM SPSS Statistics 26.

## Results

A total of 10 Bama miniature pigs were used, with 1 death attributed to puncture site bleeding and secondary infection. Severe carotid artery stenosis models (stenosis degree ≥70%) were successfully established in all pigs. The novel carotid artery stents were implanted successfully in all the right carotid arteries of the 10 pigs. Among them, 3 pigs were euthanized immediately after stent implantation for tissue sampling. One pig died 5 days after the stent implantation procedure. The remaining 6 pigs were maintained for 5 months before follow-up OCT, B ultrasound, angiography and subsequent euthanasia for tissue sampling.

### Short-term and long-term outcomes

Angiography showed the degree of the carotid artery stenosis before stenting was 84± 9%. The stenosis degree was significantly improved immediately after stent implantation with residual stenosis <5% (p < 0.001). Follow-up angiography at 5 months postoperatively showed no significant in-stent restenosis in all 6 stenotic carotid arteries.

**Figure 2.**
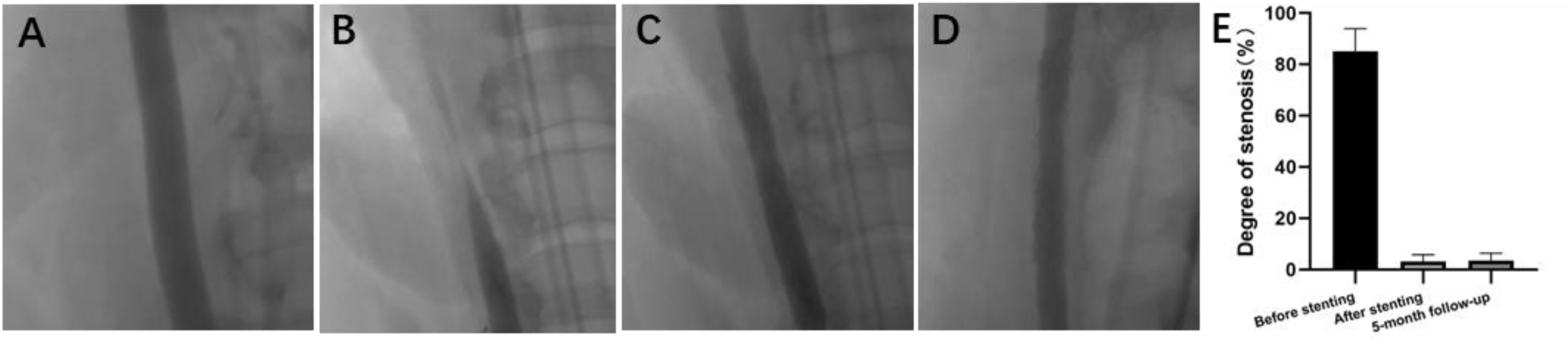
A. The normal carotid artery angiography. B. The stenotic carotid artery angiography. C. Immediate angiography after carotid stent implantation. D. Follow-up angiography 5 months after stenting. E. The bar chart demonstrates a significant improvement in the degree of carotid stenosis both immediately and 5 months after carotid stent implantation versus preoperatively.

Doppler ultrasound demonstrated immediate significant improvement in the degree of stenosis after stent implantation in all 6 stenotic carotid arteries, with no significant in-stent restenosis observed at 5-month follow-up in the 6 stenotic carotid arteries.

**Figure 3.**
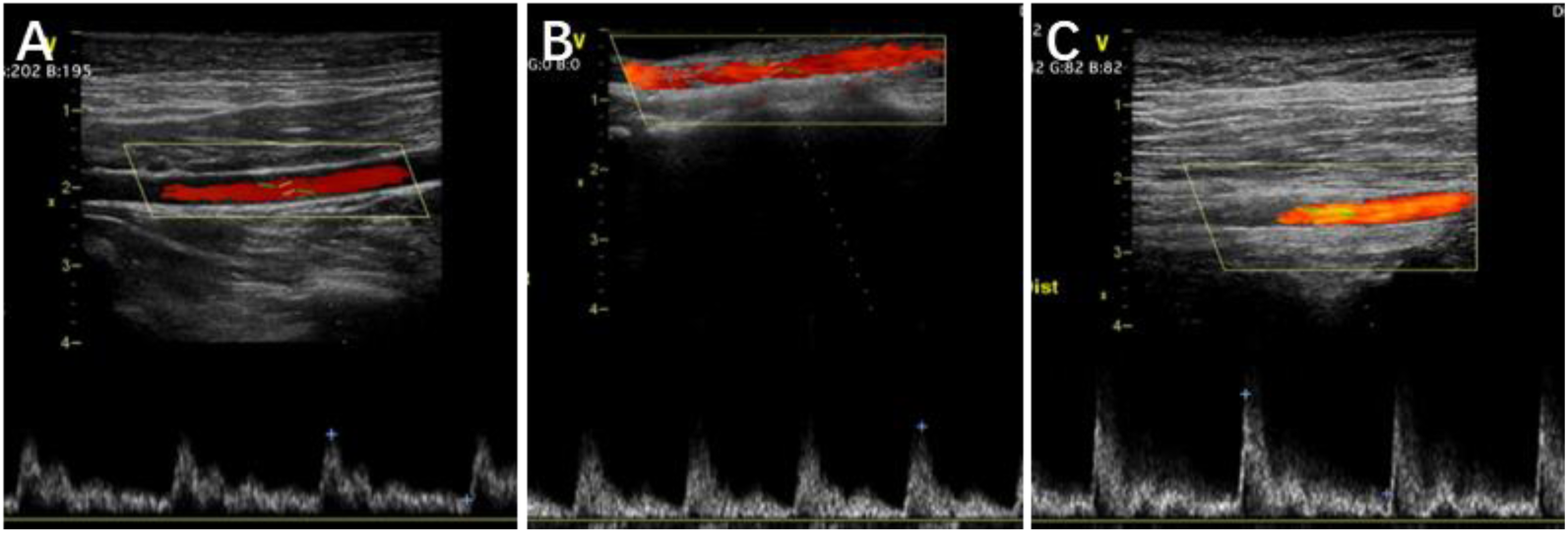
A. The normal carotid artery Doppler ultrasound. B. The Doppler ultrasound immediately after stenting for stenotic carotid artery. C. Follow-up Doppler ultrasound 5 months after stenting.

OCT showed immediate significant improvement in the degree of stenosis after stent implantation in all 6 stenotic carotid arteries, with no significant in-stent restenosis and intimal hyperplasia observed at the 5-month follow-up examination in the 6 stenotic carotid arteries.

**Figure 4.**
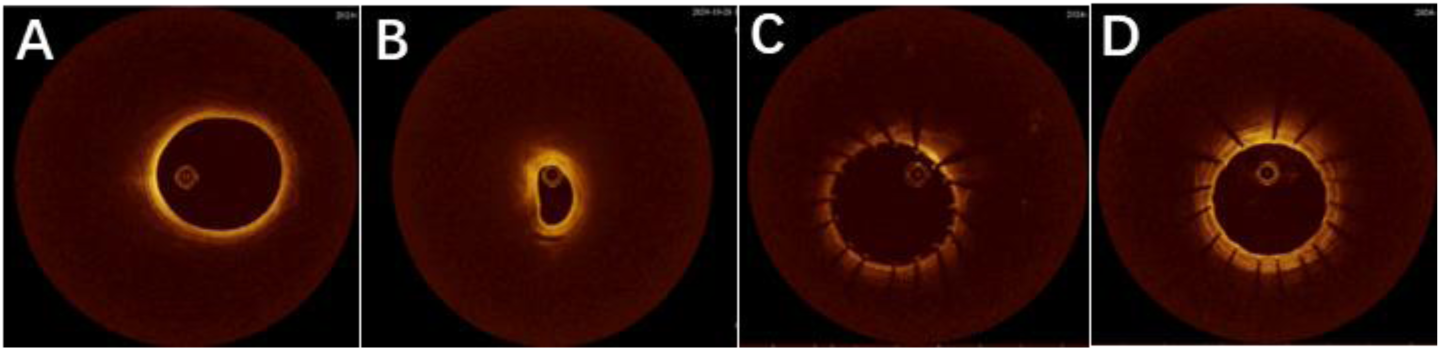
A. The normal carotid artery OCT manifestation. B. The stenotic carotid artery OCT manifestation. C. Immediate OCT manifestation after stenotic carotid stent implantation. D. Follow-up OCT 5 months after stenting showed no obvious intimal hyperplasia and in-stent restenosis.

HE staining showed significant improvement in the degree of stenosis and no obvious intimal injury immediately after stent implantation in 3 stenotic carotid arteries. And no significant in-stent restenosis or obvious intimal hyperplasia were observed at the 5-month follow-up in the 6 stenotic carotid arteries.

**Figure 5.**
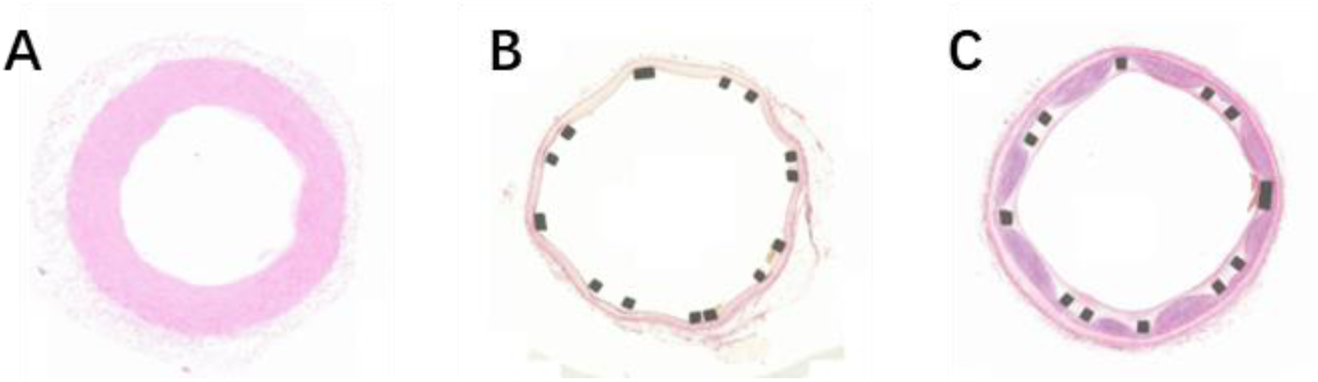
A. The normal carotid artery HE staining. B. HE staining of the stenotic carotid artery immediately after stent implantation. C. HE staining of the carotid artery 5 months after stenting showed no obvious intimal hyperplasia and in-stent restenosis.

### Complications

Except for 1 pig that died due to puncture site bleeding and secondary infection, all other pigs survived well. No complications including stent implantation failure, poor stent implantation position, blood vessel rupture, cerebral hemorrhage, cerebral infarction, in-stent restenosis, or stent migration occurred.

## Discussion

Through collaboration with Peking University’s School of Engineering and Shanghai Bochang Medical Technology Co., Ltd., our research team developed a 3-in-1 novel carotid stent system integrating balloon dilation, stent deployment, and EPD retrieval into a single device. This study demonstrated its safety and efficacy through animal experiments. Procedural and technical success was 100% and the degree of carotid artery stenosis was significantly improved after stenting.

Although CAS has been increasingly used in patients with severe carotid artery stenosis and most randomized trials comparing CAS with carotid endarterectomy (CEA) showed no significant differences in the 30-day risk of composite major adverse events like death, stroke, and myocardial infarction^9^, the risk of minor stroke has been consistently higher with CAS^10^. Various strategies have been proposed for reducing the risk of minor stroke and it’s generally admitted that the longer and more complex the procedure, the higher the risk of adverse events. Currently, there are five carotid stents available in China: Precise (Cordis)^11^, Protege^12^, Acculink (Abbott)^13^, Wallstent (Boston Scientific)^14^, and Xact (Abbott). While these five stents share similar design and manufacturing principles, none can address the two clinical challenges mentioned earlier, namely the cumbersome operation and the difficulty in retrieving the EPD.

The advantages of this 3-in-1 novel carotid stent system include: 1. Integration of EPD retrieval sheath, balloon, and stent into a single device allows sequential completion of balloon dilation, stent deployment, and EPD retrieval through stepwise advancement from proximal to distal, simplifying the procedure, minimizing the number of catheter exchanges, avoiding vascular wall injury from multiple device exchanges, significantly reducing procedure time and X-ray exposure, and minimizing adverse effects. 2. The EPD retrieval catheter remains distal to the carotid stent, enabling direct retrieval at any time, reducing retrieval difficulty and avoiding interference between the retrieval catheter and stent. 3. The “three-in-one” design reduces device consumption during procedures, saving costs for patients. In addition, this product is independently developed in China and possesses complete intellectual property rights. After successful R&D and mass production, it will significantly reduce the price of stents, decrease reliance on foreign brands, and save domestic healthcare expenditures. This study will play a positive role in promoting the localization of carotid artery stents, further reducing the occurrence of complications associated with carotid artery stenting, and reducing medical expenses for both patients and the nation.

At present, there are very few other similar studies. For example, William et al. designed a novel carotid stent system named Neuroguard IEP system with an integrated embolic filter and post-dilation balloon to treat clinically significant carotid artery stenosis and results demonstrated that the Neuroguard IEP system was safe and feasible. Ralf et al. developed the Paladin System, a novel angioplasty balloon with an integrated embolic protection filter designed to increase embolic protection during post-dilation and results showed that use of the Paladin System for post-stent dilation during CAS was safe and may effectively decrease the number of embolic particles reaching the brain, which may reduce the risk of procedure-related stroke. The novel carotid artery stent system in our study is the first 3-in-1 system which integrate balloon, stent, and EPD retrieval catheter into one system. Additionally, it features the most convenient operational workflow and requires the minimal number of catheter exchanges during carotid artery stent implantation currently.

The carotid stenosis model used in this study was originally created by the researchers, named the “fat compression method,” where autologous fat is used to externally compress the carotid artery to induce stenosis, wrapped and fixed with sterile glove material. Current literature commonly employs repeated balloon injury methods to establish carotid stenosis models^15^, but these have extremely low success rates and difficulty achieving severe stenosis^16^. Since carotid stenting generally requires at least severe stenosis as an indication^17^, such modeling methods cannot meet our study requirements. Although our fat compression method cannot fully replicate clinical atherosclerosis-induced carotid stenosis (e.g., unable to reproduce calcified lesions), the primary study objectives were to evaluate stent safety, efficacy, and stability - including smoothness of deployment, sufficient lateral and radial force to improve severe stenosis, whether it is prone to arterial rupture and bleeding, stent recoil, and intimal hyperplasia leading to restenosis - for which this modeling method is sufficient and suitable for the study’s objectives. Additionally, although the repeated balloon injury method was not used, a high-fat and high-cholesterol diet was adopted to maximize risk factors for in-stent restenosis.

Furthermore, this study has some limitations. In addition to the unconventional carotid stenosis modeling method, the study lacked strictly designed control groups. No comparisons were made with existing commercial carotid stents. At present, it is still a preliminary animal experiment, and rigorous controlled trial and human clinical trials are still required.

## Conclusion

The novel carotid stent system which integrates multiple device functions including balloon, stent, and EPD retrieval catheter, demonstrated good safety and efficacy in carotid artery stent implantation for severe carotid artery stenosis in Bama miniature pigs.

## Data Availability

All imaging and histology datasets that support the findings of this study are available from the corresponding authors upon reasonable request.

